# Ataxia as a presenting manifestation of COVID -19: Report of a single case

**DOI:** 10.1101/2020.05.24.20103648

**Authors:** Debaleena Mukherjee, Peyalee Sarkar, Souvik Dubey, Biman Kanti Ray, Alak Pandit, Durjoy Lahiri

## Abstract

Even though various neurological presentations of COVID-19 have surfaced up, ataxia as a presenting feature has rarely been reported so far. We hereby describe a confirmed case of SARS-CoV-2 infection which not only presented with ataxia but also had delayed onset of typical respiratory features. This case represents an atypical manifestation of COVID-19.

## INTRODUCTION

With the passage of time, several extra-pulmonary manifestations of COVID-19 are being reported across the globe; among which central nervous system involvement is now seemingly a well known feature of this ailment. Although several neurological features have come to attention so far, ataxia has rarely been encountered by the treating physicians. We hereby describe a confirmed case of SARS-CoV-2 infection with initial presentation as cerebellar ataxia. Noteworthy, in our patient the typical respiratory features were delayed by days which makes this case all the more important to be brought to attention.

## CASE REPORT

A 53 year old diabetic and hypertensive male patient presented to us on 14^th^ of April, 2020 with slurring of speech and difficulty in maintaining balance while walking as well as sitting since four days. He did not complain of any limb weakness, sensory symptoms or dizziness. He had a holocranial, dull aching, headache without nausea,vomiting or photophobia. No history suggestive of cranial nerve involvement, alteration of sensorium, convulsion or sphincter complaints was available. Two days later he developed shortness of breath without fever, cough or sore throat. At admission he was conscious (Glasgow Coma Scale E4,V5,M6) and afebrile. Nervous system examination revealed presence of meningeal signs including neck stiffness, Kernig’s and Brudzinski’s. Cranial nerves, motor and sensory examination were within normal limits. Cerebellar examination revealed dysarthria, appendicular and truncal ataxia. Respiratory system examination revealed tachypnoea (respiratory rate was 24 / minute, bed side peripheral oxygen capillary saturation was 90%) with bilateral vesicular breath sounds. However patient deteriorated progressively after a few hours, with fever, progressive hypoxia and drowsiness. His investigations showed a normal CT brain (Figure 1), complete blood count and metabolic parameters. Chest X ray showed heterogenous opacities in both lung fields (Figure 2). Arterial blood gas analysis revealed Type I respiratory failure. In view of deteriorating condition of patient MRI brain and lumbar puncture study could not be done. Nasopharyngeal and oropharyngeal swab sent for SARS-CoV2 Reverse Transcriptase – Polymerase Chain Reaction later came positive. He was managed with intravenous antibiotics and other supportive therapy. However patient succumbed a few hours later despite all efforts.

**Figure 1:**
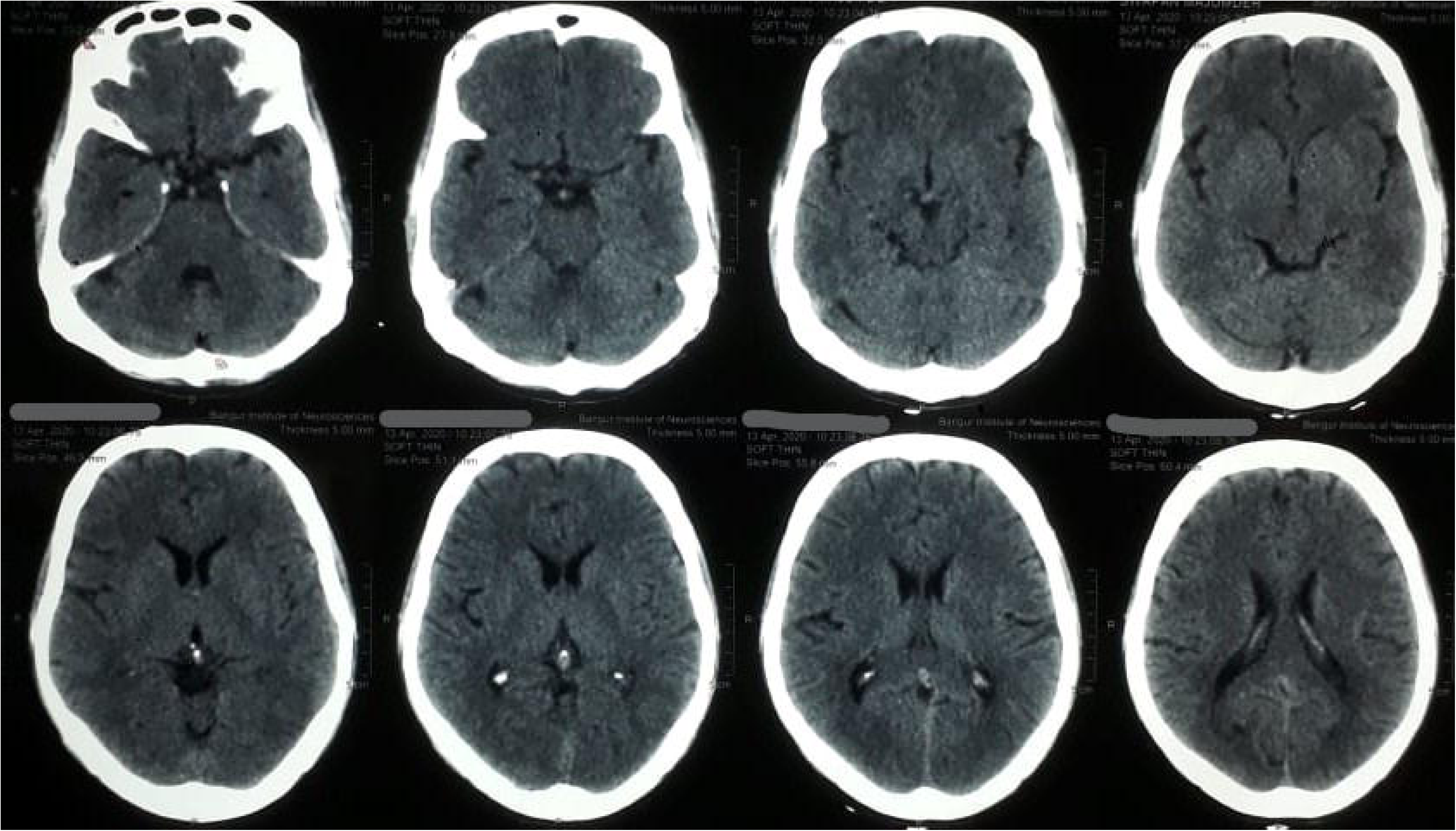

**Figure 2:**
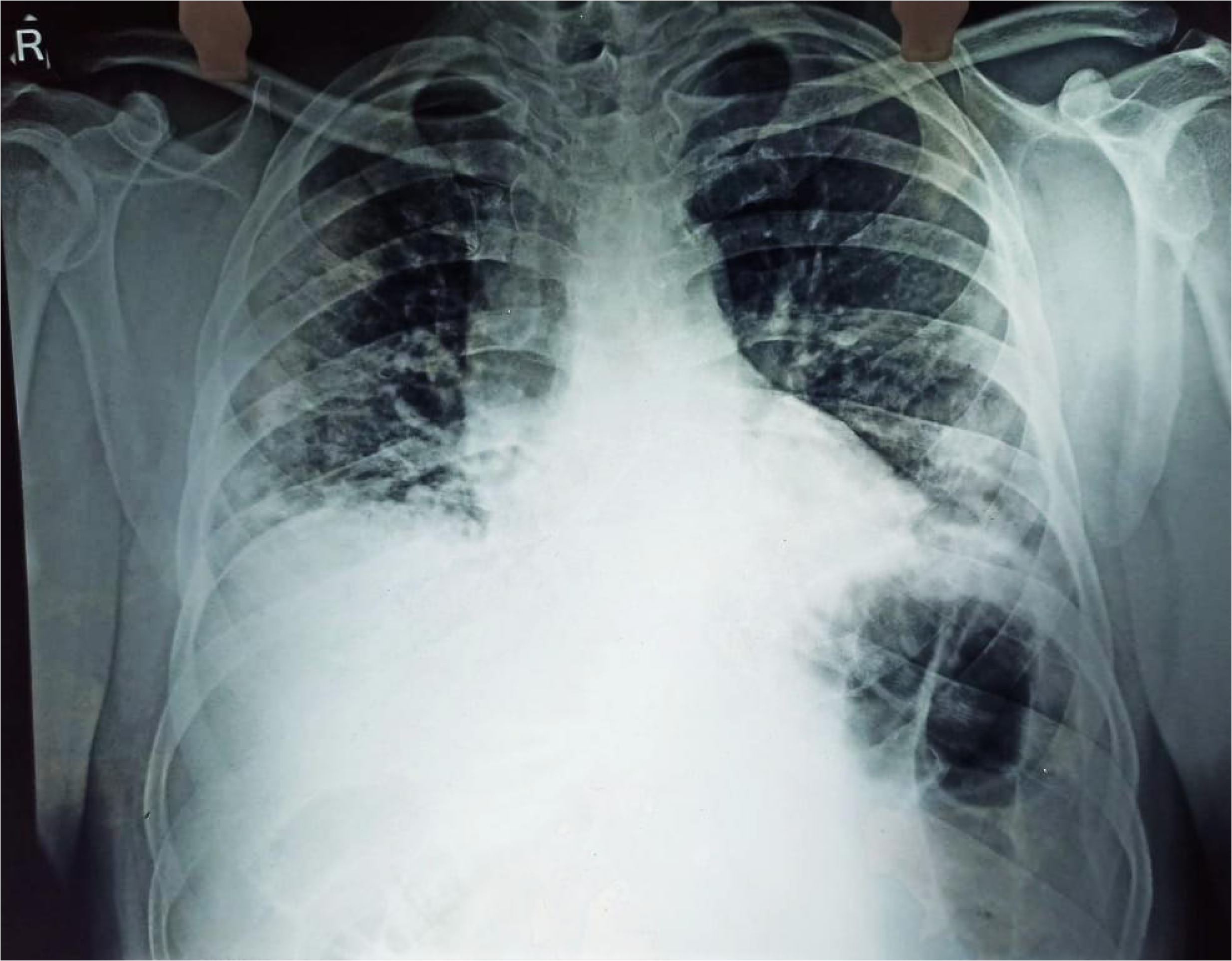

## DISCUSSION

SARS-CoV-2 virus is widely known to principally target the respiratory system. However there is growing evidence of its neuro-invasive potential. The viral particle can gain access to the CNS through both hematogenous and retrograde neuronal routes.^1^ Following a nasal infection it can also enter the CNS via the cribriform plate and ethmoid bone reaching the olfactory bulb.^2^ The virus’s affinity for ACE-2 receptors and its distribution in the nervous system also has possible contribution.^3^

Our patient presented first with acute onset neurological illness while typical features including fever and respiratory distress appeared later. Patient’s dysarthria, truncal and appendicular ataxia without any limb weakness, sensory or vestibular signs pointed towards diffuse cerebellar involvement which could be due to an infectious cerebellitis or parainfectious acute disseminated encephalomyelitis. CT brain was normal. MRI brain and lumbar puncture could not be done owing to the patient’s deteriorating condition. In the retrospective case series from Wuhan only a single case of ataxia was reported, but its anatomical localization was not described.^1^ Another recent report described ataxia in a COVID-19 patient with Miller Fisher syndrome.^4^ To the best of our knowledge ataxia remains an uncommon presentation of COVID-19 and cerebellum as its contributory anatomical substrate has not been described previously.

While hypoxia due to respiratory failure could contribute to the patient’s encephalopathy, presence of meningeal signs was a definitive evidence in favor of concomitant meningitis. However, simultaneous presence of encephalitis could not be confirmed as MRI or CSF study could not be performed. The Wuhan study reported 7.5 % percent patients presenting with impaired consciousness, however meningitis and encephalitis were not described separately.^1^ Interestingly meningoencephalitis was described in a patient who tested negative for SARS-CoV-2 in the nasopharyngeal specimen but it was detected in the CSF, emphasizing on the virus’s neuroinvasive properties.^5^ Recently a presumptive case of acute hemorrhagic necrotizing encephalopathy in COVID-19 was reported. ^6^ Another case of encephalitis was also described in a patient with fever and shortness of breath who later developed altered sensorium.

The striking part of our case was ataxia as the presenting feature followed in quick succession by meningitis and encephalopathy. Moreover our patient had simultaneous involvement of both the intra and extra axial spaces in brain. Our case exemplified an atypical clinical trajectory with neurological manifestation preceding the more well known features such as fever and respiratory distress by days. Cerebellar ataxia and meningitis remain uncommon presentations of COVID-19 and can have an unusual temporal relation with the pulmonary involvement such as in this case. Thus recognizing such neurological features as part of the clinical spectrum of COVID-19, particularly at presentation, is crucial for timely diagnosis and thereby preventing horizontal transmission.

## CONCLUSION

In sum, our case represents an atypical presentation of COVID-19. While cerebellar ataxia was the initial feature, meningitis as well as encephalopathy rapidly set in. Importantly, respiratory features had a delayed onset which made the clinical picture even more confusing. High index of suspicion on part of the clinician remains the key to solving such cases amidst the ongoing pandemic. Admittedly, we lost our patient to this deadly infection and hence would want the information to be disseminated so that many other lives might be saved.

## Data Availability

The data will not be available openly. However, on request to the corresponding author, it may be made available.

